# DISTRIBUTED LAG MODELS FOR ESTIMATING ACUTE EFFECTS OF MIXED ENVIRONMENTAL EXPOSURES IN THE CASE-CROSSOVER DESIGN

**DOI:** 10.1101/2025.10.30.25339173

**Authors:** Linda Amoafo, Amanda Bakian, Yizhen Xu, Yue Zhang

## Abstract

We present a Bayesian modeling framework designed to estimate the immediate effects of combined environmental exposures on suicide risk within a case-crossover design. Our method addresses a limitation observed in current distributed lag modeling approaches for multiple environmental exposures, which primarily focus on cohort or case-control data rather than case-crossover design. We utilize sparsity-enforcing spike-and-slab priors for variable selection, allowing the identification of significant exposures linked to the health outcome. To address clustered observations, we integrate random effects into the model. Additionally, we enhance computational efficiency and reduce dimensionality by implementing cubic polynomial reduction on the distributed lag surface. In a simulation study comparing our dimension reduction approach with a method estimating full model parameters without dimension reduction, we evaluated two referent schemes (unidirectional and bidirectional). The results demonstrate that our strategy, incorporating dimension reduction, outperforms full model parameter estimation in terms of false discovery rate, power, and mean squared error. We applied our framework to real-world data examining the association between a mixture of ambient environmental exposures and suicide risk in Utah.

## 1. Introduction

Research examining the complex relationship between environmental exposures, such as air pollution, and acute health outcomes has garnered considerable attention in recent years (Bakian et al., 2015; Pothirat et al., 2019; Go et al., 2024). Conventionally, these studies often focus on the impact of individual exposures in isolation by considering the exposure of one component at a time. Yet, individuals are exposed to multiple components in the environment simultaneously, creating a situation where assessing the adverse effects of environmental exposures independently may potentially underestimate the true health consequences that the environment entails through additive and synergistic effects (Braun et al., 2016). This limitation highlights the need for analytical approaches that aim to disentangle the individual from the joint effects of components within a mixture. Estimation bias can also arise from models that assess the exposure effect at a single time point or cumulative average over a specific exposure window. Doing so ignores the distribution pattern of the exposure series, which may potentially capture inaccurately the distributed lag nature of the exposure-response relationship (Bose et al., 2017; Leon Hsu et al., 2015; Lee et al., 2018), or inaccurately identify the time window of susceptibility to the environmental mixture (Wilson et al., 2017a). The distributed lag model (DLM) addresses this issue by studying the relationship between health outcomes and exposures summarized at regular intervals over a given time frame. The DLM also enables researchers to examine critical exposure windows, i.e., crucial time frames of susceptibility during which adverse health outcomes are influenced by environmental exposures.

The application of DLMs to assess the influence of exposures on health outcomes has been extensively studied (Schwartz, 2000; Zanobetti et al., 2000). A primary area of recent research on DLMs is to address the problem of significant autocorrelation in the exposure measure. Various techniques have been introduced, including smoothing splines, principal component analysis, and Gaussian processes, to achieve a smooth distributed lag surface (Warren et al., 2012, 2016, 2020; Chang et al., 2015; Wilson et al., 2017b; Gasparrini et al., 2017). Other study focus in DLM include the following: nonlinear exposure-response estimation (Gasparrini, Armstrong and Kenward, 2010; Gasparrini, 2011; Gasparrini et al., 2017; Mork and Wilson, 2022), two-step estimation approach which examines additive models and the correlation between multiple exposures at different time points (Warren et al., 2013; Chen, Mukherjee and Berrocal, 2019; Antonelli, Wilson and Coull, 2021), variable selection among multiple exposures by incorporating Bayesian shrinkage priors (Antonelli, Wilson and Coull, 2021), as well as the identification of critical exposure windows (Warren et al., 2020; Andrews and Mallows, 1974). Some approaches have also employed Bayesian estimation (Warren et al., 2016; Wilson et al., 2017b, 2021; Antonelli, Wilson and Coull, 2021), along with kernel machine regression (Wilson et al., 2022).

For the study of acute exposures with rapid onset health effects, particularly for rapidly occurring outcomes such as asthma exacerbation, myocardial infarction, and suicide death, the case-crossover design is considered suitable (Maclure, 1991). The case-crossover design may be preferable under certain conditions to other study designs commonly used to investigate exposure-outcome associations such as cohort and case-control studies. For example, cohort studies can be expensive and have limited statistical power for rare outcomes (Setia, 2016) and case-control studies may suffer from bias due to retrospective exposure assessment and control group selection (Maclure, 1991). In terms of identifying representative controls, the case-control designs select control individuals, whereas the case-crossover designs select control time windows. Estimation bias can arise in case-control study designs when the control sample is not representative of the case sample. On the other hand, the case-crossover design contrasts an individual’s exposure history during a specific “case” period—when the outcome of interest occurs—with the exposure during “control” periods—when the outcome does not occur (Maclure, 1991). The main advantage of the case-crossover design is that it eliminates the necessity of controlling for time-invariant confounding factors, such as age, sex, and underlying medical conditions. This is because, instead of being compared to the exposure history of another individual, each person’s exposure history is only compared to their own baseline, which is the time period immediately preceding the outcome event. As a result, the effect of the exposure of interest can be more easily isolated, and its estimate can be more accurately estimated.

However, the existing methods for DLM do not account for the impact of clustered observations, which is inherent in the study design as the case window is compared to the control windows for the same individual. To address this challenge, we adopt the method by Antonelli, Wilson and Coull (2021) for a mixed-effects model to account for subject-specific heterogeneity. The approach relies on a Bayesian mixed-effects modeling framework to estimate the acute effects of mixed environmental risk factors on an acute health outcome. We use random effects to account for clustering in the data resulting from observations collected on the same subject. Additionally, we employ spike-and-slab priors on the distributed lag curves to detect significant exposures associated with the outcome.

We demonstrate the applied use of our approach by examining the acute effect of a mixture of ambient environmental exposures on the risk of suicide mortality. Prior research has shown that exposure to a range of environmental pollutants, such as particulate matter and ozone, as well as meteorological factors, such as solar radiation and increasing temperature, might adversely affect the risk of suicidal behavior (Ragguett et al., 2017; Davoudi et al., 2021). Understanding the specific impact of mixed environmental exposures on the risk of suicide remains a complex challenge. Understanding the specific impact of mixed environmental exposures on suicide risk is a complex challenge, and our goal is to enhance this understanding using our proposed approach.

The remainder of the paper is organized as follows: Section 2 introduces the proposal by describing the Bayesian inference framework. Section 3 presents results from a simulation study comparing the unidirectional and bidirectional referent schemes. In Section 4, we apply our proposed method to real data to explore the effects of air pollution and meteorological exposures on suicide, followed by a discussion and conclusion in Section 5.

## 2. Method

### 2.1. Notation

We consider the following variables for our case-crossover design. Let *Y*_*ik*_ ∈ {0, 1} indicate the presence or absence of a certain outcome in the *k*th case or control window for person *i*, where *i* = 1, …, *n* and *k* = 1, …, *K*. Without loss of generality, we let *k* = 1 represent the case window and *k* = 2…, *K* be the control windows. We let *Z*_*ijkt*_ be the exposure of the person *i* to the *j*th pollutant at the lagged time point *t* within the *k*th case/control window, where *j* = 1, …, *p* and *t* = 1, … *T*. Additionally, we have a vector of observed covariates denoted as *X*_*ik*_ ∈ ℝ^*q*^ during the *k*th time window for person *i*. The observed data for person *i* across the *K* time windows, denoted as *O*_*i*_, is formatted as (*Y*_*i*_, *Z*_*i*1_, …, *Z*_*ip*_, *X*_*i*_), which equals

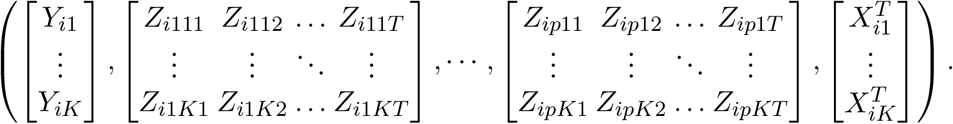

We write (*Z*_*i*1_, …, *Z*_*ip*_) as matrix *Z*_*i*_ and (*X*_*i*1_, …, *X*_*iK*_)^*T*^ as *X*_*i*_. The observed dataset consists of *n* samples {*O*_1_, …, *O*_*n*_}, and we assume that these samples are independent and identically distributed.

### 2.2. Model

To examine the effects of the mixture of exposure variables on the outcome, conditional on the design matrix *X*_*i*_, we adopt a distributed lag model (DLM) framework for the binary outcome variable *Y*_*ik*_. The probability of an event for subject *i* with observed exposure matrix *Z*_*i*_ is denoted as *π*_*ik*_. The model is specified as follows:

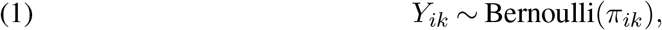

where *π*_*ik*_ is defined as:

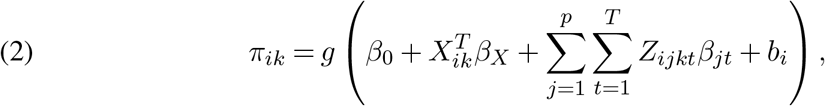

with *b*_*i*_ representing the individual random intercept for each subject. Exposure coefficients *β*_*jt*_ are assumed to be shared across time windows, which are indexed by *k*. Here, the function *g*(.) corresponds to the link function, which dictates the relationship between the linear predictor and the probabilities. Under probit and logistic regression models, *g*(.) is defined as the Gaussian cumulative distribution function and the inverse expit function, respectively.

### 2.3. Estimation and Variable Selection

In this study, we use a Bayesian approach for parameter estimation, employing sparsity-enforcing priors known as spike and slab priors on the regression coefficients for variable selection. The spike and slab priors were originally introduced by Mitchell and Beauchamp (1988) for Bayesian variable selection in standard linear regression models. We apply variable selection on the exposures; for each exposure, a spike and slab latent variable is shared among the corresponding set of lagged exposure statuses. This results in a mixture between a point mass at zero (spike) and a multivariate normal distribution (slab) for the prior distributions of the regression coefficients for exposures. The formulation of the priors is as follows. Let *β*_*j*_ = (*β*_*j*1_, …, *β*_*jT*_) represent the vector of the regression coefficients associated with the *j*th exposure, *Z*_*ij*_. We specify the spike and slab prior for *β*_*j*_ as

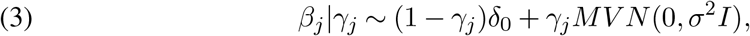

where *γ*_*j*_ is a latent variable indicating whether the exposure variable *j* has a substantial effect on the outcome. When *γ*_*j*_ = 0, it implies that *β*_*j*_ = 0, effectively removing the entire effect of *jth* exposure from the model. Additionally, we assume the following prior for the binary indicator variable *γ*_*j*_:

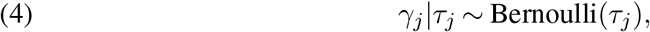

where *τ*_*j*_ is a parameter determining the probability of *γ*_*j*_ being 1, reflecting the significance of *j*th exposure. Other priors used in the model include:

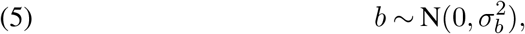

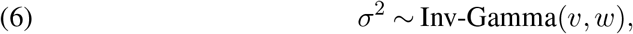

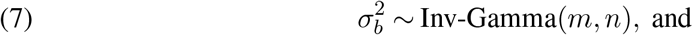

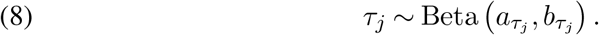

Let *θ* = {*β*_0_, *β*_*X*_, *β*_*j*_, *τ*_*j*_, *σ, σ*_*b*_ |*j* = 1, …, *p*}, representing all parameters in the above framework and having prior distribution denoted as *θ ∼ p* (*θ*). Then, the posterior distribution for the Bayesian binary model is given by

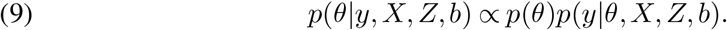

We rely on the Markov Chain Monte Carlo (MCMC) algorithm for posterior computation to estimate the parameters and perform variable selection. All parameters are directly sampled from their conditional posterior distributions using Gibbs sampling. We implemented all computations using JAGS (Martyn Plummer, 2023) under the logistic link function, considering that it would be easier to interpret the model with the estimated coefficients being odds ratio. Because the posterior distributions under the logistic link function do not have a close analytic form, we provide the derivation of posterior distributions for each model parameter under the probit link function in Supplementary A. In the computation, we set initial values for each parameter to a random sample from its respective prior distribution; we burn in half of our posterior samples and thin for every 3rd sample. MCMC convergence is examined using trace plots with multiple chain evaluation and a convergence diagnostic statistic proposed by Gelman and Rubin (1992). Inference is based on random samples from the posterior distributions. For instance, the posterior inclusion probability for each exposure variable can be calculated as follows:

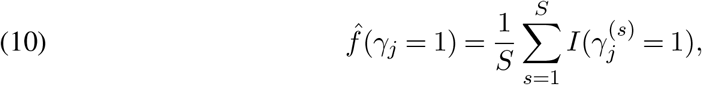

where *S* denotes the number of posterior samples. Subsequently, exposures are considered important to be included if their posterior inclusion probability exceeds a specified threshold (e.g., 0.5) (Li et al., 2022).

### 2.4. Dimension Reduction

Parameter estimation becomes challenging because of the high dimensionality when *p* or *T* is large, as the cardinality of the distributed lag parameters {*β*_*jt*_ | *j* = 1, …, *p, t* = 1, …, *T*} is *pT*. Our approach aims to represent the distributed lag parameters using weighted combinations of basis functions denoted by 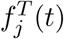 to minimize the parameter space dimension while maximizing flexibility in estimating the surface effects. We propose to reduce the parameter space dimension using principal component analysis or cubic polynomial reduction by imposing smoothness conditions on the distributed lag curves across time within a particular exposure. Instead of allowing *T* different parameters for each exposure, we let there be *C*_*j*_ number of basis parameters that retain as much flexibility in the resulting surface effect estimates as possible, where *C*_*j*_ ≤ *T*. We define the following,

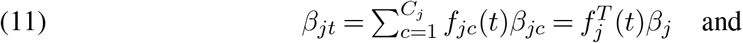

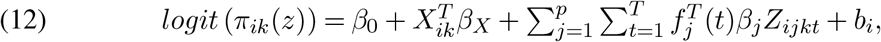

where 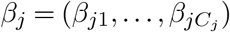.

In our simulation and data application, we used cubic polynomial splines for dimension reduction. We note that principal component analysis (PCA) can also be adopted for a similar purpose (Antonelli, Wilson and Coull, 2021) by choosing the number of basis functions, *C*_*j*_, to explain a specific fraction of the data’s variability. Dimension reduction in DLMs reduces the data’s complexity while preserving the lagged relationship’s key features. Details for dimension reduction schemes are included in Supplementary B.

### 2.5. Identification of Critical Exposure Lag Windows

While we focus on identifying exposures associated with the outcome, we also want to determine critical windows of susceptibility. When an exposure exhibits a critical window, it is more likely to be included in the model due to the selection of hyperparameters from *γ*. However, we still need a method to determine which periods are most strongly linked to the outcome. Following the approach in Antonelli, Wilson and Coull (2021), we address this by utilizing the Bayes-p measure to evaluate the presence of critical windows. Specifically, Bayes-p for exposure *j* at the *t*th time window is defined as

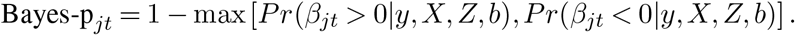

A Bayes-p value close to 0 indicates evidence of a critical window at the *t*th time window. The identification of critical windows is based on the criterion that Bayes-p_*jt*_ *<* 0.025, which aligns with the concept of credible intervals proposed by Warren et al. (2020), where significance is indicated by having the 95% posterior credible interval not including zero.

## 3. Simulation Study

We adopt the simulation setting in Antonelli, Wilson and Coull (2021) to access the performance of our proposed environmental mixture distributed lag model with case-crossover design.

### 3.1. Data Generating Mechanism

For the simulation, we assume 3 exposures and 7 time points, i.e., *p* = 3 and *T* = 7. The *p* exposures are generated using an autoregressive model, allowing for correlation among and within exposures across time. Each subject’s exposure values over time are drawn according to the following equation:

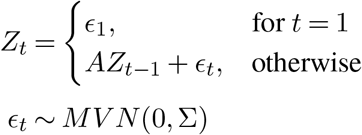

where *Z*_*t*_ represents the p-dimensional vector of exposures at time *t*. The p-dimensional matrix A is a diagonal matrix with 0.95 on the diagonals, inducing temporal correlation within each exposure over time. We allow correlation among exposures by specifying 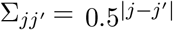. We assume the first two exposures to have nonzero main effects, where the first exposure has a quadratic effect over time and the second exposure has a constant effect over time. The functional form of the exposure effect is illustrated in Figure 1. The outcome at a *k*th time window, *Y*_*ik*_, is generated using the Bernoulli distribution as described in equation (2) conditional on blocks of the previous *T* time points of the *p* exposures.

**Fig 1.**
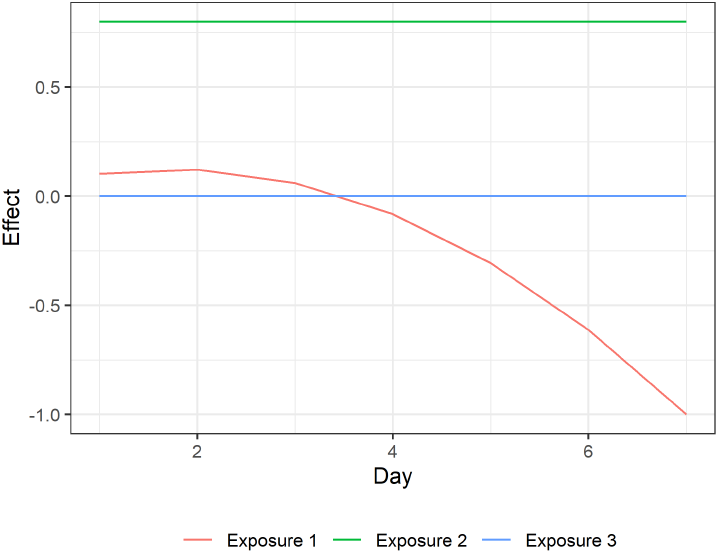
True exposure effect of distributed lag surface for each exposure at t=1,2,…,7

#### 3.1.1. Referent Selection

We explored two different referent schemes in our simulation process: the unidirectional and the bidirectional referent schemes. We conducted our analysis by considering up to three control window selections for each referent scheme. For the unidirectional referent scheme, we included all three blocks of previous *T* time points if the subject had at least three blocks available. However, if the subject had fewer than three blocks of previous T time points, we included as many blocks as were available. In contrast, the bidirectional referent scheme required the subject to have at least three blocks of both previous and subsequent *T* time points. If a subject did not meet this criterion, we included the number of time points *T* that were available for both the preceding and subsequent time points. To evaluate the performance of each referent strategy, we considered two scenarios. In the first scenario, estimation was performed with dimension reduction on the distributed lag surface using the cubic polynomial. The second scenario involved estimation at all lag days without any dimension reduction (full estimation). We considered sample sizes of 100, 200, 300, 400, 500, and 1000 in our analysis. For each combination of the referent scheme and estimation scenario, we generated a total of 500 simulated samples.

### 3.2. Performance Measures

We assess the performance of these approaches using various metrics, including power, false discovery inclusion probability, 95% pointwise credible interval coverage, and mean squared error (MSE) (Antonelli, Wilson and Coull, 2021). Power is the average probability of including important effects in the model, calculated by averaging the posterior inclusion probabilities of all important exposures across all simulations. False discovery inclusion probability represents the average posterior inclusion probability of null effects in the model, determined by averaging all null exposures across all simulations. The coverage of the 95% point-wise interval measures the proportion of simulations in which the 95% credible interval of the distributed lag curves contains the true parameters, averaged over all important parameters in the model. Lastly, mean squared error is the average squared difference between the posterior mean and the true values for each parameter, averaging all exposures, time points, and simulations.

### 3.3. Results

Figure 2 illustrates the results of our simulation study by showing the evaluated performance metrics of the method with and without dimension reduction. The false discovery rate (FDR) consistently favors the method with dimension reduction, showing lower rates across all sample sizes. This indicates that the variable selection performance is significantly improved when employing dimension reduction estimation compared to the full estimation. Moreover, the posterior inclusion probabilities exhibit a notable difference between the two methods. The full estimation tends to produce estimates with slightly higher probabilities of inclusion, particularly as the sample size increases. Consequently, the power of both methods remains comparable, but it improves with larger sample sizes.

**Fig 2.**
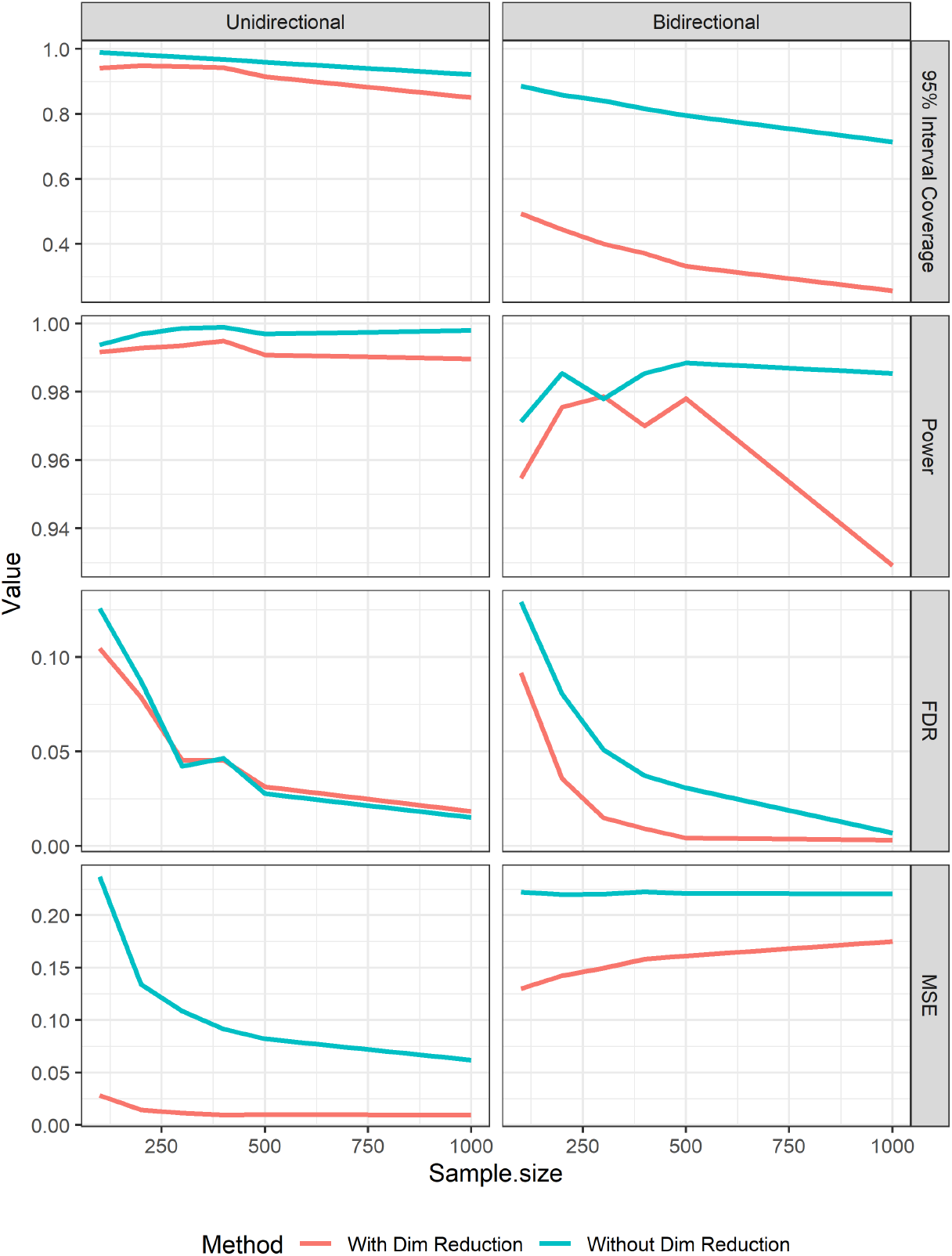
Simulation study results for the performance measures. column1 (Unidirectional Referent Scheme), column2 (Bidirectional Referent Scheme), row1 (95% Interval coverage), row2(Power), row3 (False discovery rate),row4 (Mean squared error)

In terms of interval estimation, the full estimation demonstrates slightly higher 95% interval coverage rates due to the wider credible intervals (as depicted in Figures 3 and 4). These wider intervals provide a greater chance of encompassing the true effects, leading to higher coverage rates than the nominal value for the full estimation. Additionally, the mean squared error (MSE) analysis underscores the superiority of the method with dimension reduction, as it consistently yields lower MSE values. This highlights the improved accuracy in estimating values for this approach, demonstrating its efficacy over the method without dimension reduction across all sample sizes.

**Fig 3.**
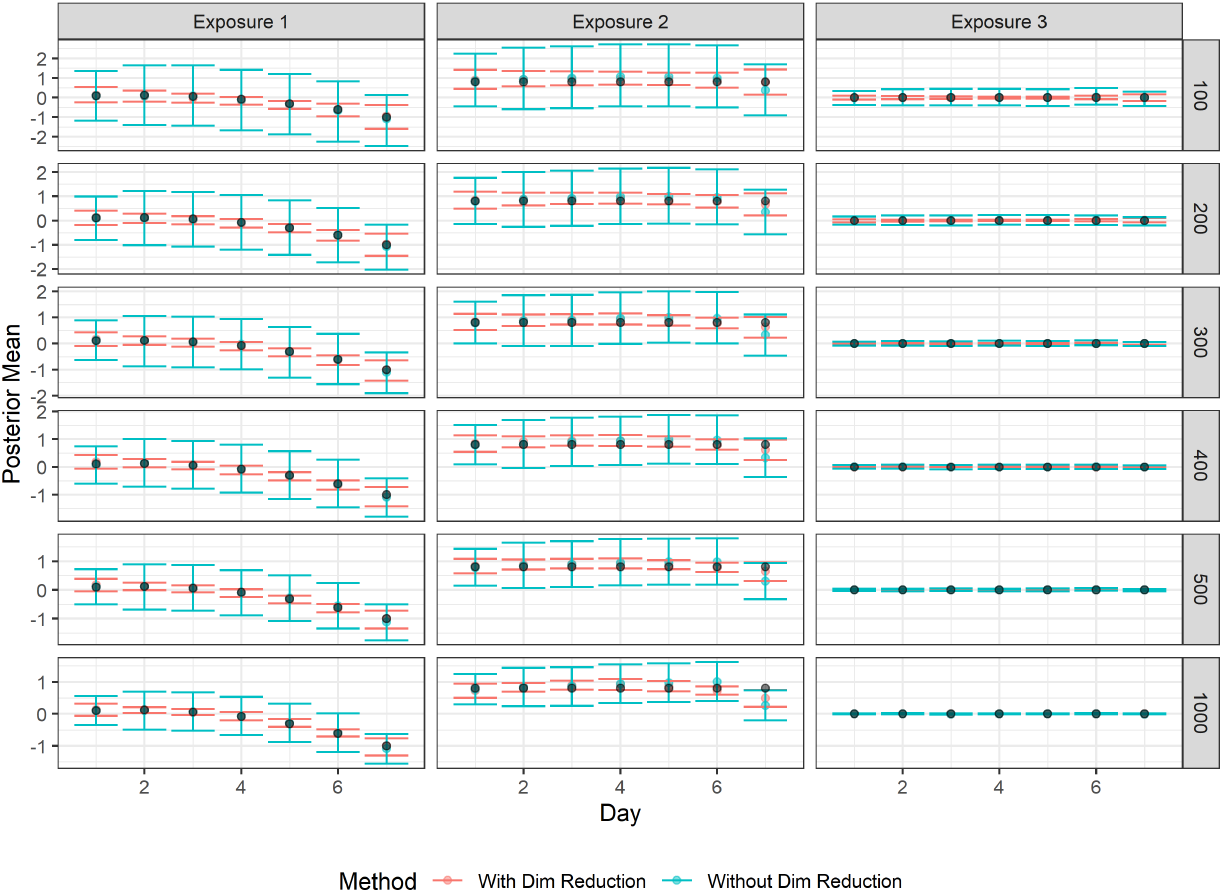
Simulation study results of the exposure-response surface estimation for unidirectional referent scheme for each sample size

**Fig 4.**
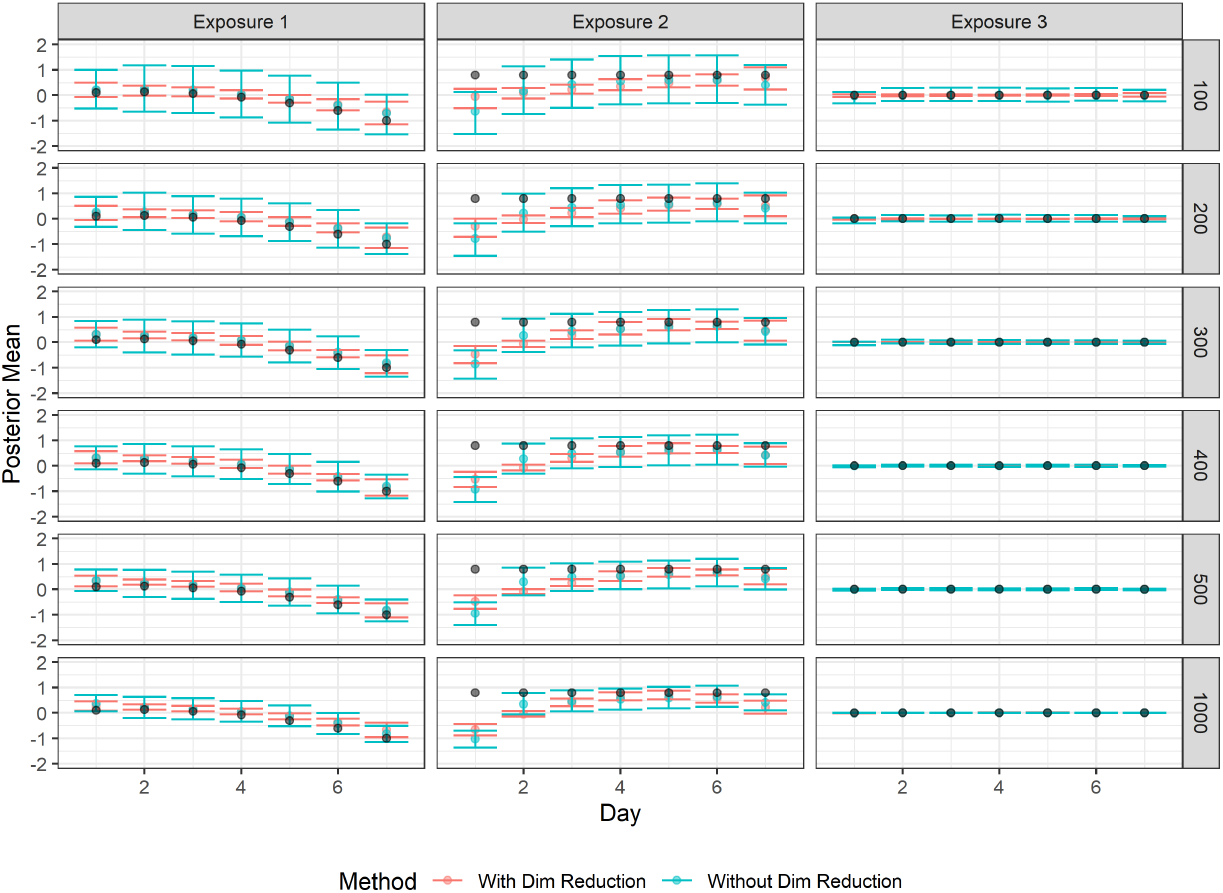
Simulation study results of the exposure-response surface estimation for bidirectional referent scheme for each sample size

Regarding the referent schemes, the unidirectional referent consistently displays better performance across almost all metrics when compared to the bidirectional referent. The bidirectional referent slightly outperforms the unidirectional referent regarding false discovery rate (FDR), where the results are remarkably comparable. However, in all other performance metrics, the unidirectional referent scheme demonstrates superior performance, showing its suitability for the specific application.

Figures 3 and 4 present the estimation of the exposure-response surface along with their corresponding 95% confidence intervals for the unidirectional and bidirectional referent schemes, respectively. Each figure illustrates the true estimate as a black dot at the center for each of the 7 days, while the point estimates, along with the 95% credible intervals averaged over all simulations, are provided for each exposure.

In the case of the unidirectional referent scheme, the exposure-response surface for exposure 1, which is quadratic, demonstrates accurate estimation. Notably, utilizing dimension reduction consistently results in narrower credible intervals at all sample sizes, whereas the method without dimension reduction produces wider credible intervals, with the width decreasing as the sample size increases. Nonetheless, each of the narrower credible intervals contains the true estimate. For exposure 2, which exhibits a flat surface, a similar conclusion can be drawn, but the method without dimension reduction indicates a slight bias in the point estimate, especially as the lag days increase.

Exposure 3, which has no discernible effect, shows consistent estimates under both methods. However, there is some false discovery with credible intervals for smaller sample sizes, but this bias diminishes as the sample size increases. On the other hand, the bidirectional referent scheme reveals notably more bias, the true estimates do not align with the point estimates. While results for exposure 3 results remain relatively similar between both referents, those for exposure 2 exhibit significantly higher bias, particularly for the 1-day and 7-day lags. This bias is due to the usage of the cubic polynomial dimension reduction, which assumes the exposure function over time is cubic. In Supplematary C, we include the simulation results for both referent schemes, which showed less biased results with linear exposure function for exposure 2 over time.

## 4. Application to a Study of the Effects of Environmental Mixtures on Suicide Mortality in Utah

We illustrate our method in analyzing the effect of seven environmental mixtures on the risk of suicide death in Utah, USA from 2000-2016. The exposures of interest include daily averaged ambient air pollutants such as particulate matter smaller than 2.5 microns in diameter *PM*_2.5_, nitrogen dioxide *N* 0_2_, and ground-level ozone *O*_3_, which were obtained from the national Harvard ensemble model (Di et al., 2017, 2019a,b), as well as daily averaged weather exposures such as relative humidity, temperature, precipitation, and solar radiation obtained from the gridMET model (Abatzoglou, 2013). Population-wide suicide deaths from 2000-2016 were identified through a long-standing collaboration with the Utah Office of the Medical Examiner as well as from death certificates (see Docherty et al. (2020); Coon et al. (2020) for more details). For the application, we use a random sample of 500 suicide events and focus on the summer season from June 9 to September 12, with an average daily high temperature above 81°F (weatherspark, 2024). In this analysis, we use cubic polynomials for dimensional reduction and the unidirectional time-stratified referent scheme. The case window was identified with the case day, which is the event day (based on the date of death), and all previous six days before the event date were included as lagged days. The control time windows for each event were identified, with the control days being all previous days falling on the same day and month as the event, and the days before the control day were included as the lagged days. The model is estimated using five MCMC chains. Each chain consists of 10,000 iterations. The first half of the iterations are discarded as burn-in, and every 5th posterior sample is retained after thinning.

The posterior inclusion probabilities are given in Table 1. While none of the air pollutants exhibited exceptionally high inclusion probabilities, weather-related factors, especially temperature and solar radiation, demonstrated notable inclusion probabilities exceeding 0.5. In Figure 5, the exposure-response relationship is illustrated for an interquartile range increase in exposures over time, accompanied by their respective 95% credible intervals. Whereas Figure 6 presents the Bayes-p values for exposures with a posterior inclusion probability ≥ 0.5 at each time-point.

**TABLE 1.** Posterior Inclusion Probabilities.

| Exposures | PIP |
| --- | --- |
| PM<sub>2.5</sub> | 0.0998 |
| NO<sub>2</sub> | 0.2414 |
| Ozone | 0.2394 |
| Precipitation | 0.2524 |
| Solar Radiation | 1.0000 |
| Maximum Temperature | 0.9984 |
| Maximum Relative humidity | 0.1400 |

**Fig 5.**
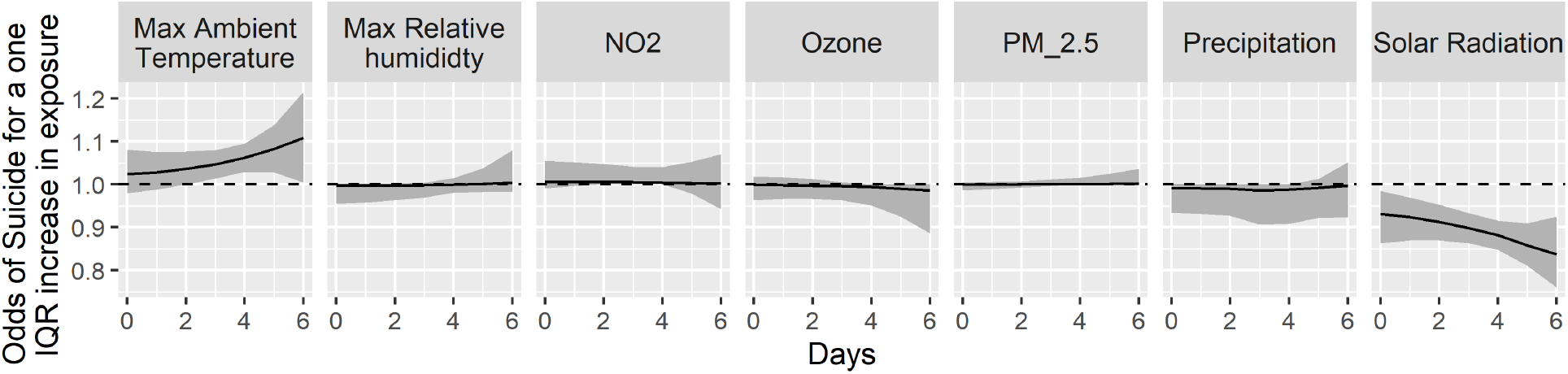
Distributed Lag results from analysis of the effect of the six environmental mixtures on suicide

**Fig 6.**
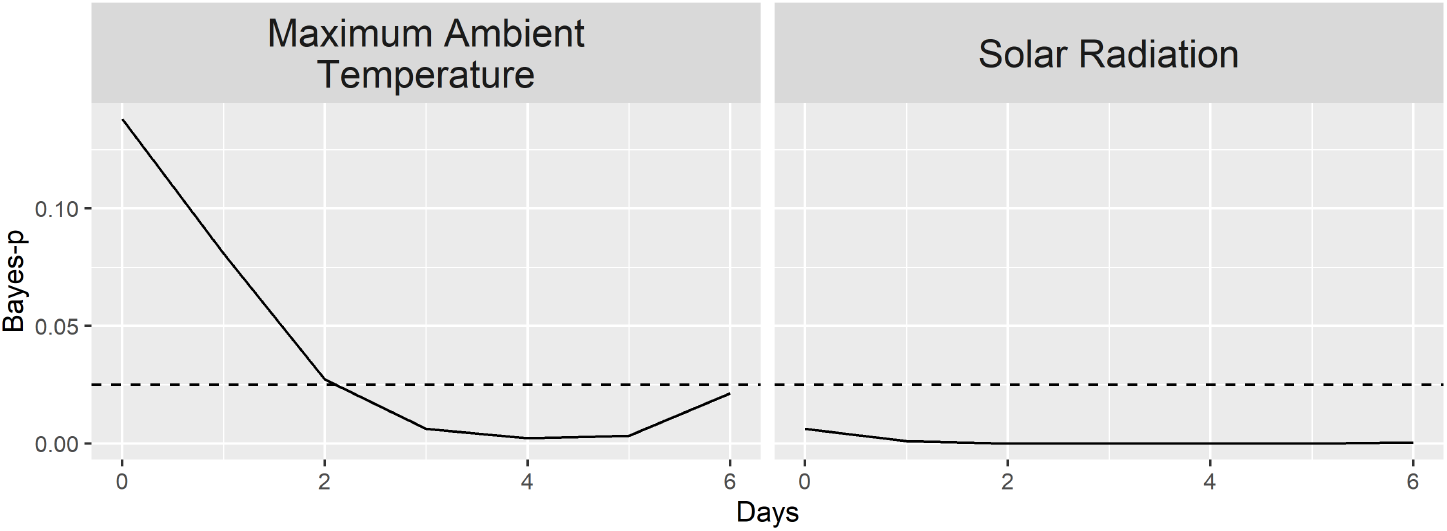
Bayes-p for each exposure on each day to assess critical window

**Fig 7.**
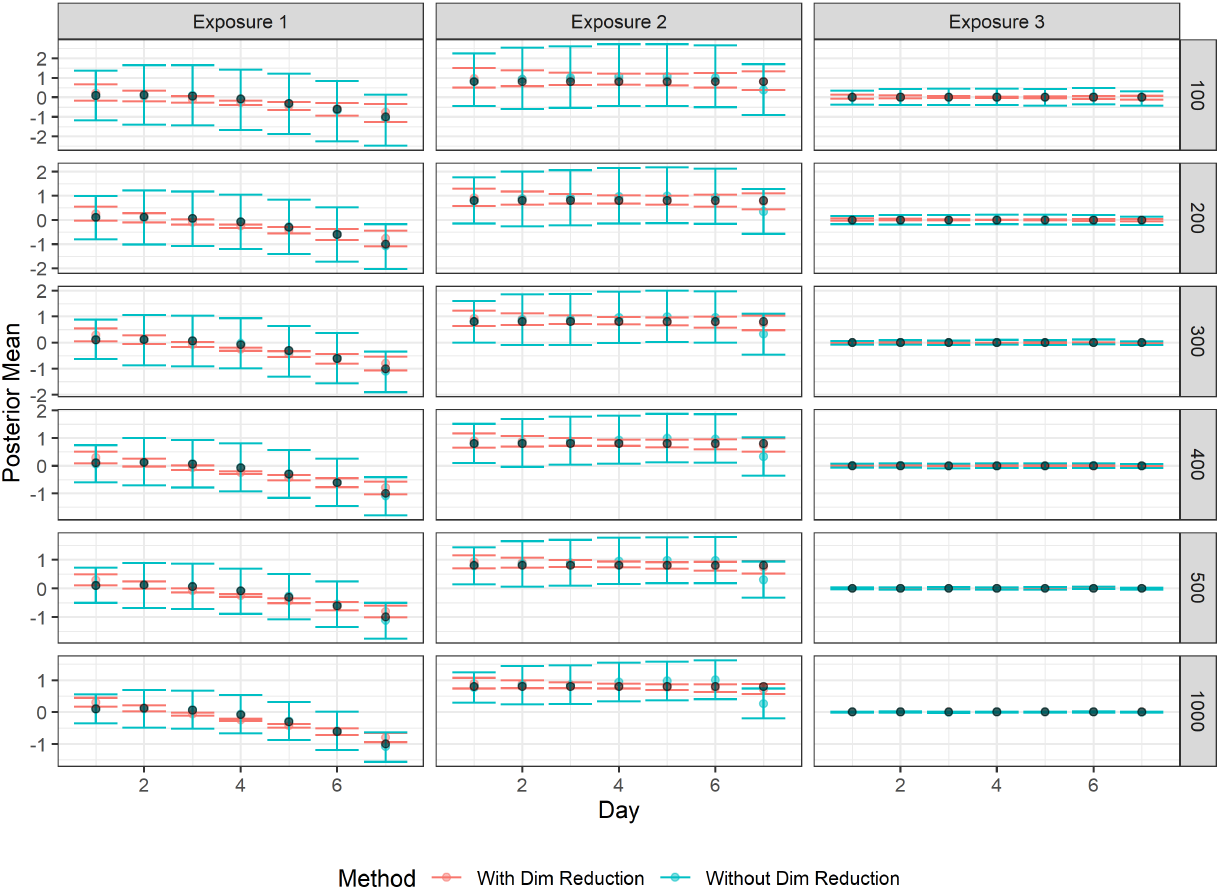
Simulation study results of the exposure-response surface estimation for unidirectional referent scheme for each sample size, applying linear dimension reduction

**Fig 8.**
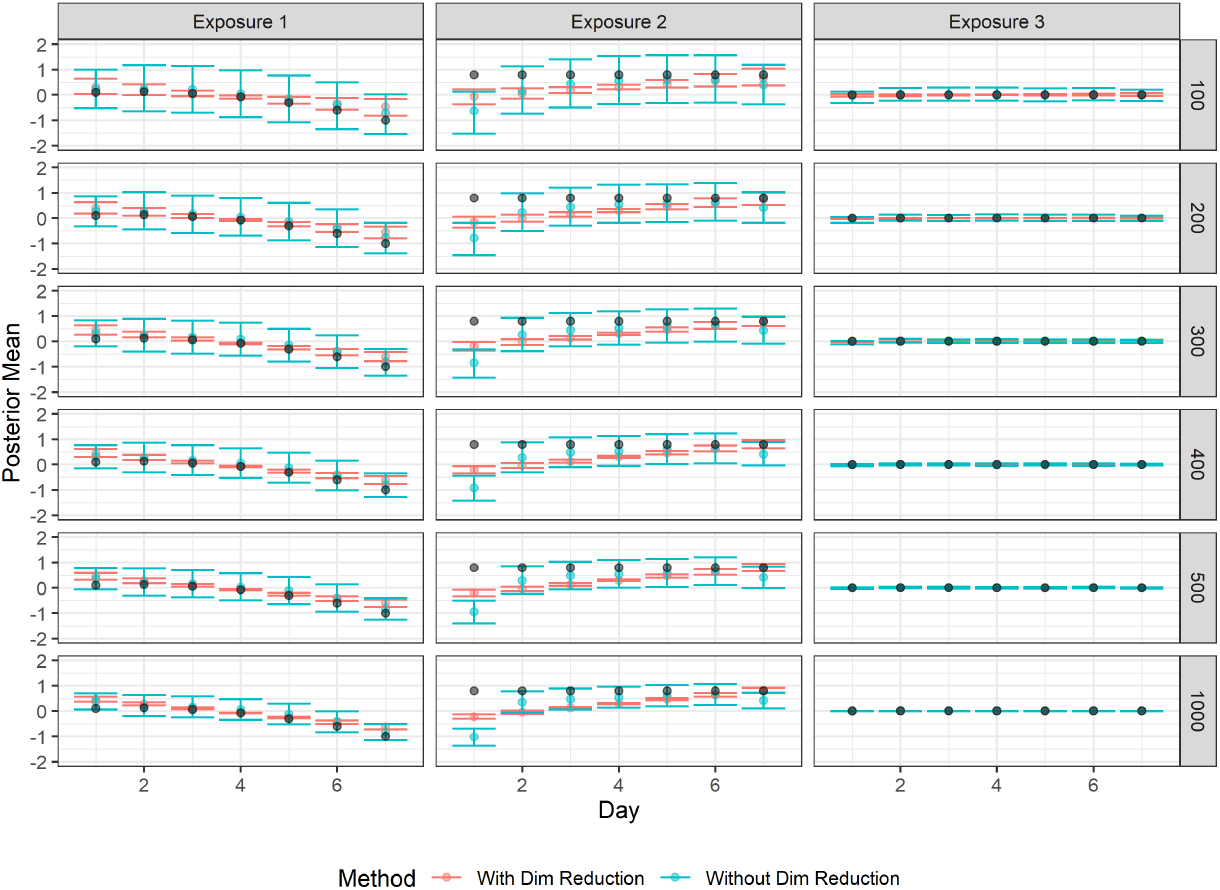
Simulation study results of the exposure-response surface estimation for bidirectional referent scheme for each sample size, applying linear dimension reduction

Based on the posterior inclusion probabilities, only solar radiation and temperature showed discernible effects, with solar radiation negatively impacting suicide rates. We observed an association between suicide and rising temperatures, indicating that as temperatures increase in the summer, suicide rates also tend to rise.

A Bayes-p value closer to 0 indicates the presence of a critical window. For solar radiation, all Bayes-p values were ≤0.025, signifying critical windows at all specified times. For temperature, the critical window spans from 3 to 5 lagged days.

## 5. Discussion and Conclusion

In this paper, we developed a distributed lag model within a case-crossover design to estimate the acute effects of mixed environmental exposures on a health outcome, specifically focusing on disentangling the impact of various ambient environmental exposures on suicide mortality risk Our approach effectively addressed the limitations of existing methods by incorporating dependencies among observations, such as accounting for the impact of the clustering effect. We utilized Bayesian methods and spike-and-slab priors to perform variable selection, allowing the identification of significant exposures associated with the outcome.

Through the simulation study, we evaluated the performance of our proposed method in comparison to the full estimation approach and examined two referent schemes: unidirectional and bidirectional. The results demonstrate the superiority of our dimension reduction approach in terms of false discovery rate, power, and mean squared error. Notably, the unidirectional referent scheme consistently outperformed the bidirectional scheme across various performance metrics, providing a crucial finding given the relevance of unidirectional referent selection for fatal outcomes like suicide.

While it is technically invalid to select referents after the case event, Lumley and Levy (2000) demonstrated that the bias from sampling referents beyond the time at risk is relatively minor compared to exclusively sampling referents before the index time in the presence of a time trend in exposure. This finding has led to the widespread adoption of bidirectional referent selection strategies among researchers conducting case-crossover analyses. However, our simulation results suggest that reducing bias comes with a cost for distributed lag models, which was not seen in the single lag models. The results indicate that when the exposure reduction function is suboptimal for a specific exposure, both referent strategies introduce bias, but the bias is more pronounced for the bidirectional than the unidirectional referent scheme. While it would be beneficial to include a method for estimating an optimal exposure reduction strategy for each exposure, this falls beyond the scope of our current paper and is left for future studies.

Our method’s ability to identify critical windows of susceptibility enhances our understanding of the temporal relationships between environmental exposures and outcomes. Applied to the study of environmental mixtures on suicide in the population of Utah, our findings highlight a significant association between weather exposures—specifically temperature and solar radiation—and suicide, in contrast to the limited evidence for an association with air pollutants. Our findings align with prior research (Burke et al., 2018; Cheng et al., 2021; Kent et al., 2009). Burke et al. (2018) and Cheng et al. (2021) reported an increased suicide risk linked to elevated temperatures. In our summer-focused setting, this effect is observed with a lag of two to six days. Additionally, Kent et al. (2009) identified a dose-response relationship between sunlight exposure and cognitive function, correlating lower sunlight levels with impaired cognitive status—a factor associated with an elevated risk of suicide. This study not only opens avenues for further exploration into critical windows of susceptibility but also sheds light on the dynamic associations between environmental factors and acute outcomes such as suicide. Furthermore, the proposed method holds promise for application in exploring other acute health outcomes, including but not limited to asthma exacerbation, pneumonia, and myocardial infarction.

## Data Availability

All data produced in the present study are available upon reasonable request to the authors

## SUPPLEMENTARY MATERIAL

We introduce a latent variable *W*_*ik*_ where each *W*_*ik*_ follows a normal distribution. The augmented probit regression model has the following hierarchical structure

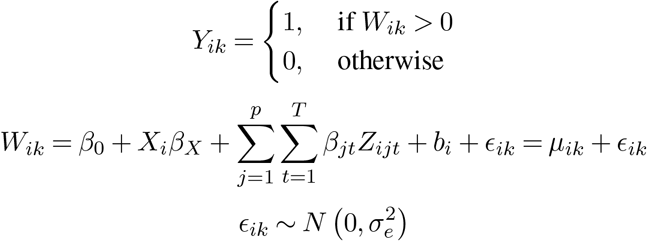

Let *θ*_−*v*_ and *D* represent all parameters except the parameter (*v*) and data, respectively. The conditional distributions of the parameters are as follows. For *τ*_*j*_

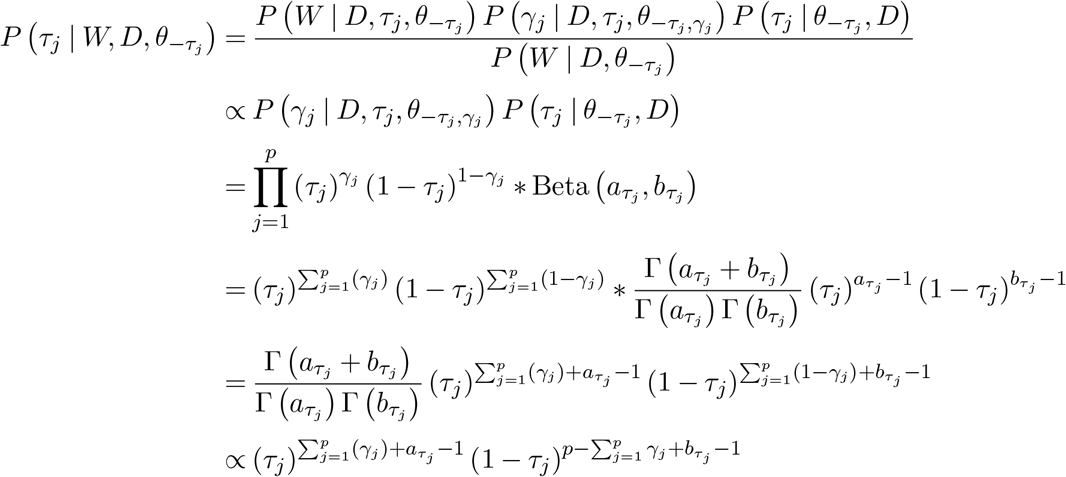

This is the kennel of Beta 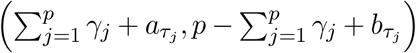, which will be the posterior distribution for *τ*_*j*_. For *γ*_*j*_, we derive its posterior distribution as follows,

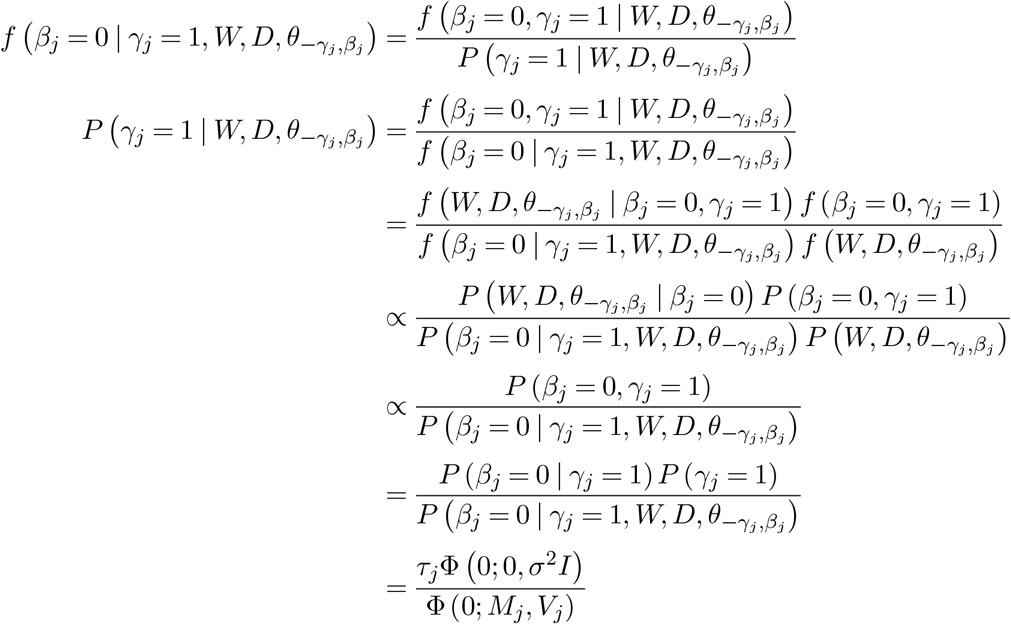

*M*_*j*_ and *V*_*j*_ are the mean and variance of the posterior distribution of *β*_*j*_ which will be derived later.

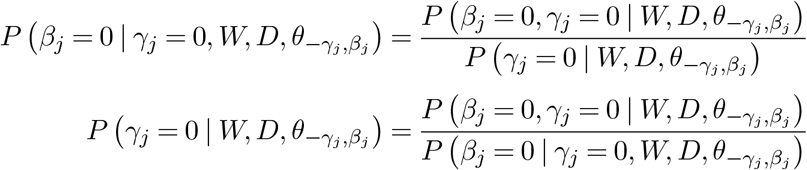

following steps from before we have

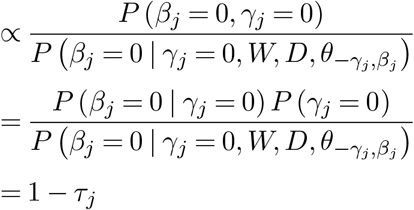

We can then sample *γ*_*j*_ from the Bernoulli distribution by normalizing the probabilities. After obtaining *γ*_*j*_ we update *β*_*j*_ and *W*_*ik*_ using the following factorization,

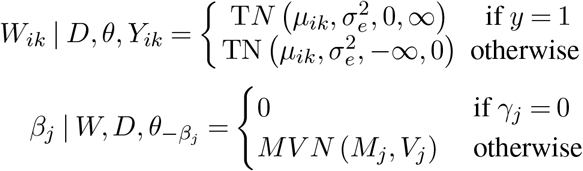

For *β*_*j*_ we have,

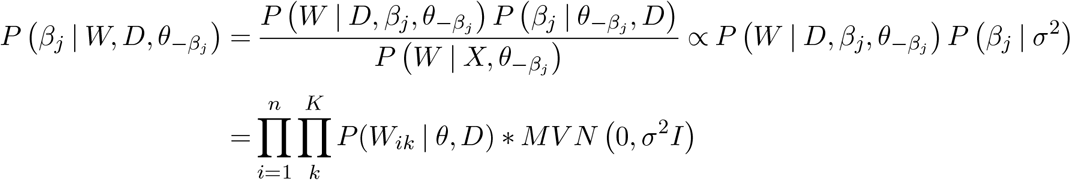

Let 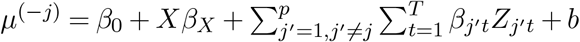 be the mean vector without exposure *j* and *Z*^(*j*)^ be the matrix for exposure *j*

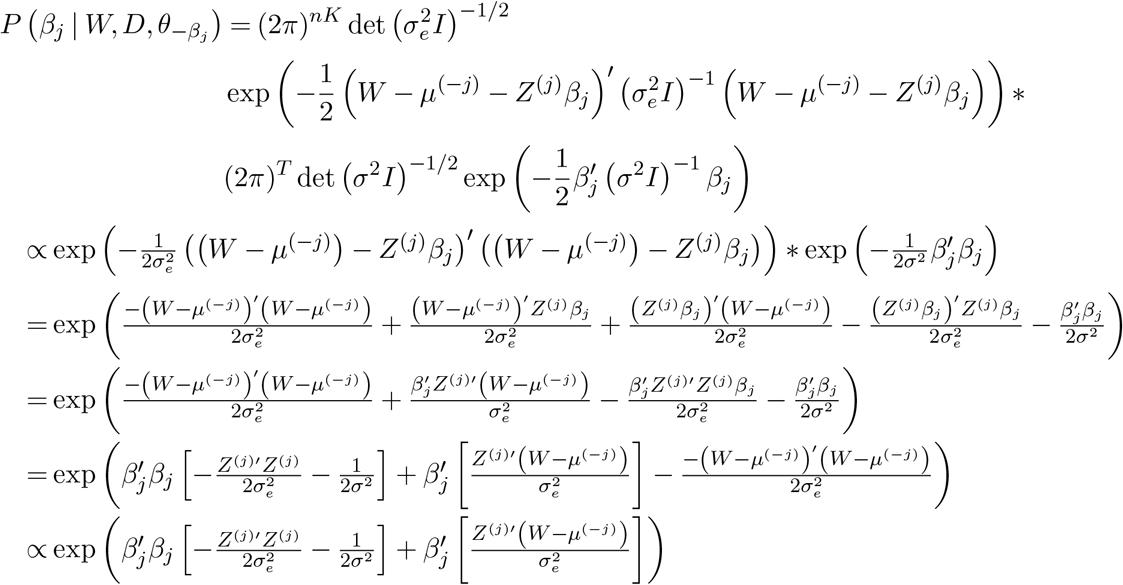

By completing the squares, we have,

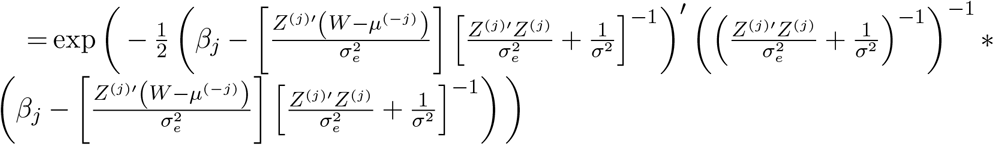

This is the kennel of a *MV N* (*M*_*j*_, *V*_*j*_) where 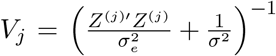 and 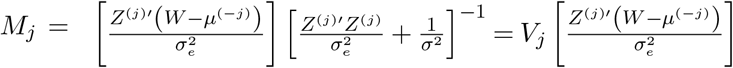 For *b* we have,

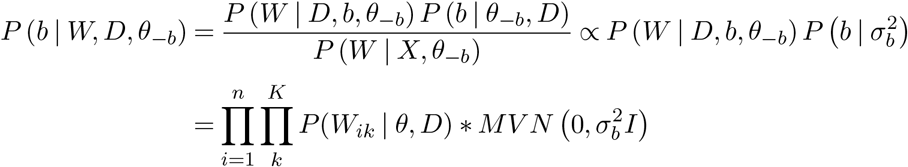

With similar derivation as we did with *β*_*j*_ we have the full conditional for *b* as

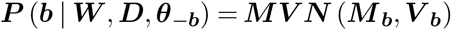

Where 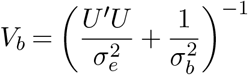 and 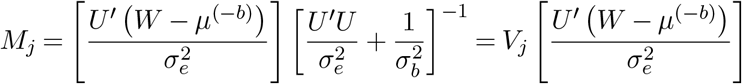

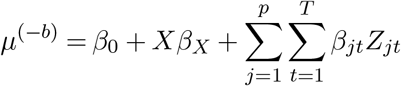

*U* is the nKxn design matrix for b with entries of 1 for person *i* and 0 for person *i*^*′*^≠ *i*. For 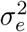 we have,

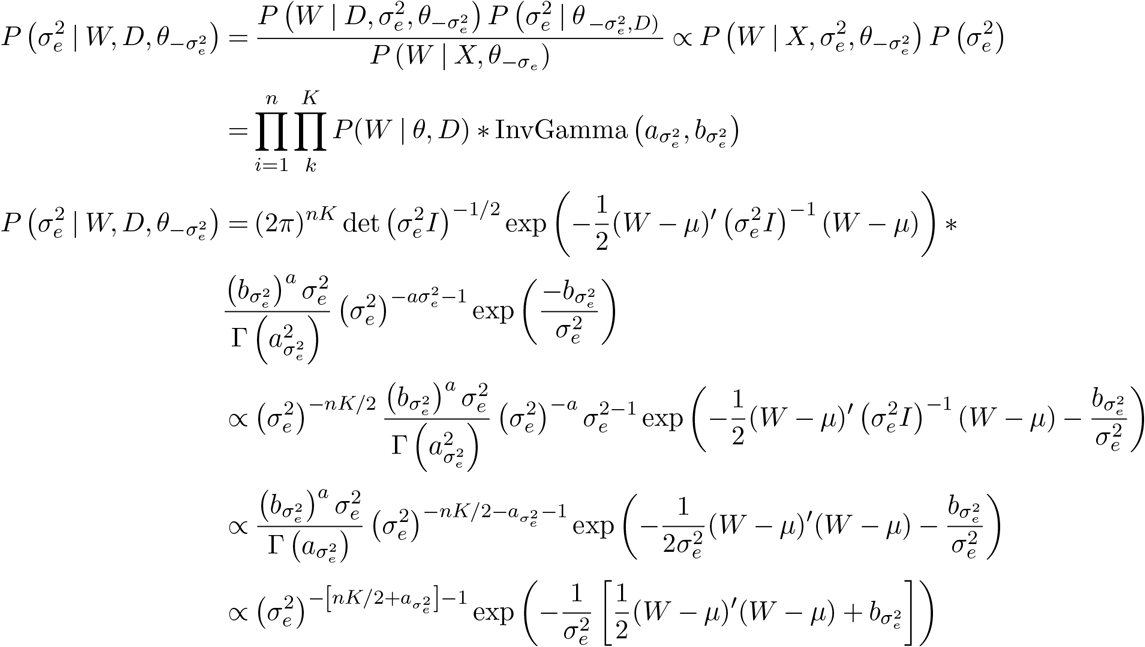

This is the kennel of InvGamma 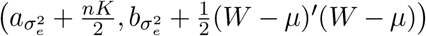

### Supplement A. Posterior Distributions under Probit Link Function

This supplementary material provides the derivation of posterior distributions for each model parameter under the probit link function.

#### BASIS FUNCTIONS

The matrix of *C*_*j*_ number of basis functions *f*_*j*_ for lagged time dimension reduction for each *j*th exposure using the PCA and cubic polynomial are given as follows;

PCA

- Calculate the principal components by deriving the eigenvectors from the covariance matrix based on lagged exposure measurements.
- Sort the eigenvalues and their corresponding eigenvectors. Let *C*_*j*_ represent the number of principal components required to explain 80% of the data’s variability.
- Form the projection matrix by selecting the top *C*_*j*_ principal components. This matrix serves as the foundation for basis functions with dimension *T × C*_*j*_ + 1, with the first column being **1** − *vector* for the constant term.

Cubic Polynomial

Let T represent the number of lagged times. The basis functions for a cubic polynomial are generated as follows:

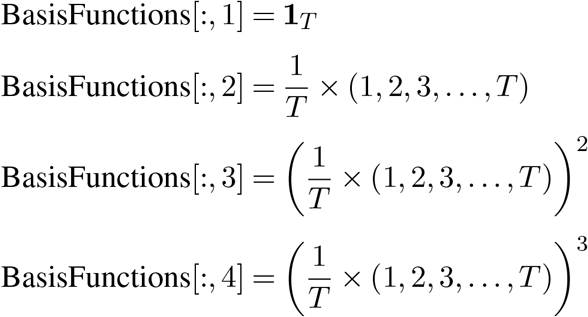

The first column is a constant term, the second column represents a linear term, the third column represents a quadratic term, and the fourth column represents a cubic term.

### Supplement B

This supplementary provides the basis functions for reducing dimension reduction on the distributed lag surface for PCA and Cubic polynomial

### Supplement C. Linear Dimension Reduction

In this supplementary, we implemented a linear transformation on the distributed lag surface to achieve dimension reduction. It is noteworthy that the bias associated with exposure 2 exhibits a reduction when compared to the application of a cubic polynomial spline for dimension reduction. This observation underscores the enhancement in fit achieved through dimension reduction, particularly when the optimal exposure-response surface function is known. Additionally, we observed that the unidirectional referent scheme

